# Airway emergency management in a pediatric hospital before and during the COVID-19 pandemic

**DOI:** 10.1101/2020.09.25.20201582

**Authors:** Christopher S Thom, Hitesh Deshmukh, Leane Soorikian, Ian Jacobs, John E Fiadjoe, Janet Lioy

**Affiliations:** Division of Neonatology, Children’s Hospital of Philadelphia, Philadelphia, PA; Division of Neonatology, Perelman School of Medicine at The University of Pennsylvania, Philadelphia, PA; Perinatal Institute, Cincinnati Children’s Hospital Medical Center, Cincinnati, OH; Division of Otolaryngology, Children’s Hospital of Philadelphia, Philadelphia, PA; Department of Anesthesiology and Critical Care, Children’s Hospital of Philadelphia, Perelman School of Medicine, University of Pennsylvania, Philadelphia, PA, USA; Department of Anesthesiology and Critical Care Medicine, Perelman School of Medicine at the University of Pennsylvania, Philadelphia, PA

**Keywords:** Pediatrics, neonatology, infant, intubation, emergencies

## Abstract

**Objective:** Children’s hospitals frequently care for infants with various life-threatening airway anomalies. Management of these infants can be challenging given unique airway anatomy and potential malformations. Airway emergency management must be immediate and precise, often demanding specialized equipment and/or expertise. We developed a Neonatal-Infant Airway Safety Program to improve medical responses, communication, equipment usage and outcomes for infants requiring emergent airway interventions.

**Patients and Methods:** All patients admitted to our quaternary neonatal and infant intensive care unit (NICU) from 2008-2019 were included in this study. Our program consisted of a multidisciplinary airway response team, pager system, and emergency equipment cart. Respiratory therapists present at each emergency event recorded specialist response times, equipment utilization, and outcomes. A multidisciplinary oversite committee reviewed each incident.

**Results:** Since 2008, there were 159 airway emergency events in our NICU (∼12 per year). Mean specialist response times decreased from 5.9±4.9 min (2008-2012, mean±SD) to 4.3±2.2 min (2016-2019, p=0.12), and the number of incidents with response times >5 min decreased from 28.8±17.8% (2008-2012) to 9.3±11.4% (2016-2019, p=0.04 by linear regression). As our program became more standardized, we noted better equipment availability and subspecialist communication. Few emergency situations (n=9, 6%) required operating room management. There were 3 patient deaths (2%).

**Conclusions:** Our airway safety program, including readily available specialists and equipment, facilitated effective resolution of airway emergencies in our NICU and multidisciplinary involvement enabled rapid and effective changes in response to COVID-19 regulations. A similar program could be implemented in other centers.

## Introduction

Unique anatomy can complicate neonate and infant airway management [1,2], and unexpected airway emergencies arise frequently in neonatal intensive care units. Further, management of neonates delivered at quaternary children’s hospitals with complex airway issues can demand “on call” multidisciplinary teams [3]. Timely airway emergency interventions are necessary and life-saving.

We developed a multidisciplinary Neonatal-Infant Emergency Airway Program to facilitate rapid airway interventions. An oversight committee reviewed all incidents. Collaboration with Otolaryngology and Anesthesia teams facilitated management of bedside airway procedures and rare cases that required operating room interventions. These emergency procedures underwent important changes in response to the COVID-19 pandemic. Herein we present our longitudinal experiences, finding specialized equipment availability and usage, subspecialist response times, and retrospective oversight to have facilitated consistently rapid interventions and optimal outcomes. As NEAR4Kids and NEAR4Neos programs have improved pediatric airway management [4,5], our findings may similarly help improve clinical airway emergency responses.

## Materials and Methods

### Patients

Patients admitted to our quaternary neonatal and infant intensive care unit (NICU) were included in this study, which was deemed exempt from oversight by the Children’s Hospital of Philadelphia Institutional Review Board.

### Methods

Our Neonatal-Infant Airway Safety Program included a multidisciplinary response team and specialized equipment (Fig. 1 and Table 1). A hospital-wide notification system alerted relevant personnel to event locations by a single pager phone button push. Once activated, respiratory therapists brought specialized equipment to bedside, including bronchoscopes, Benjamin scopes, fiberoptic scopes, a variety of endotracheal tubes and sizes, laryngeal mask airways (LMAs), and surgical airway equipment (Table 1). Nurses retrieved emergency code carts, including intravenous access equipment, medications, and other supplies. Minimum personnel at each event included an Attending Neonatologist, respiratory therapist, nurse, an ENT and/or Anesthesiologist. Video laryngoscopy is the standard of care for all intubations in our NICU.

**Table 1.** Personnel and equipment present and available at all Airway Emergency Responses.

|  |
| --- |
| **Personnel** |
| Neonatology – Attending and Fellow Physicians, Nurse Practitioners, Nursing Staff |
| Otolaryngology – Attending, Fellow, and Resident Physicians |
| Anesthesia – Attending, Fellow, and Resident Physicians |
| Respiratory therapists |
| **Airway Equipment** |
| Direct laryngoscopy handles and blades (size 00, 0, 1) |
| Video laryngoscopy handles, blades, and monitors (size 0, 1) |
| Benjamin laryngoscopes |
| Flexible bronchoscopes (sizes 2.2, and 2.8 with suction) and monitor tower |
| Endotracheal Tubes (2.0, 2.5, 3.0, 3.5, 4.0) |
| Alligator forceps (large, small) |
| Tracheostomy surgical set with neonatal and pediatric size tracheostomy tubes |
| Laryngeal mask airways (traditional and intubating LMAs, sizes 1, 1.5) |
| Nasopharyngeal airways (sizes 6.5-8.5) |
| **Other equipment** |
| Hospital-wide notification system and phones |
| Personal protective equipment (gowns, gloves, N95 masks, surgical masks) |
| Viral filters for in-line use (15 mm inner diameter, 22 mm outer diameter) |

**Fig. 1.**
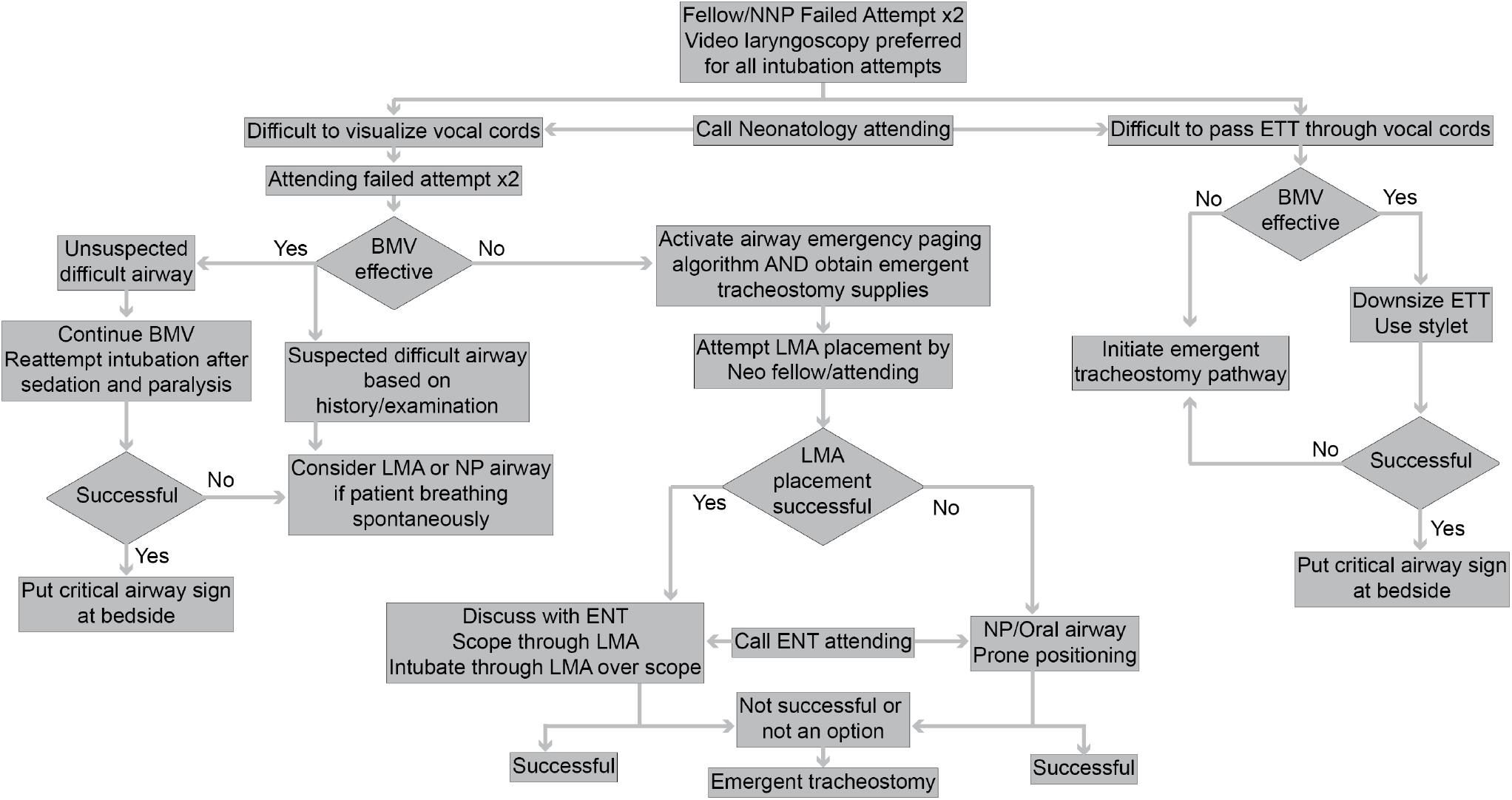
Neonatal Airway Emergency Response algorithm flow sheet. Initial steps assume management by neonatology team, including Fellow, Neonatal Nurse Practitioner (NNP) and/or Neonatology Attending physician. Video laryngoscopy was the standard of care for intubation attempts. If initial attempts failed, an emergency response page was sent to Otolaryngology (ENT) and Anesthesia teams. LMA, laryngeal mask airway. NP, nasopharyngeal airway. BMV, bag mask ventilation. ETT, endotracheal tube.

In response to COVID-19, we also made available a separate cart equipped with extra N95 masks, eye protection, gloves, hand sanitizer, and gowns for all responding personnel. All personnel donned full personal protective equipment (PPE), since airway interventions were considered aerosol generating procedures. Rapid COVID-19 tests were performed for all new admissions for airway issues, as well as semi-elective diagnostic airway procedures, such as nasopharyngeal laryngoscopy. Patients were presumed positive until negative testing was confirmed.

Respiratory therapists or nurses present during each event recorded response times, communication, equipment utilization and outcome after each event on Emergency Tracking Forms. Events and forms were analyzed by a multidisciplinary Airway Safety Monitoring team, comprised of NICU, ENT and Anesthesia physicians, as well as respiratory therapists, to address clinical complications or systemic problems. Given event infrequency and severity, we believe we have recorded all events over the past decade. Any events missed were likely during initial months of program establishment, as event records are now routine in our NICU.

## Results

We separated and analyzed our findings into three epochs to monitor statistical trends. Initial data was collected for these events from 2008-2012. From 2013-2015, standardized measures were enacted and procedures became streamlined. Since 2016, our airway emergency response program has been well established. The number of events has remained constant at ∼12 per year (Fig. 2A). Patients managed in the course of these events varied widely in sizes and diagnoses.

**Fig. 2.**
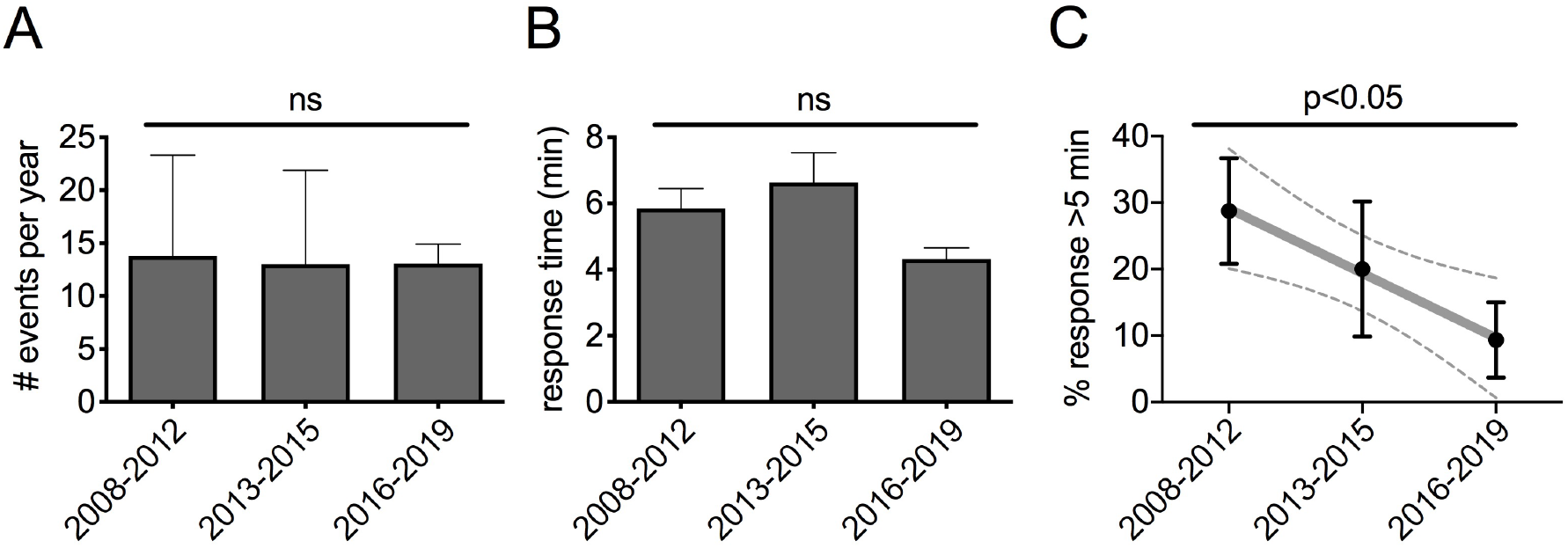
Airway Emergency Response events and response times over the study period. A. The number of Airway Emergency Response events per year over the study duration. ns, no significant difference (p=0.98 by one-way ANOVA). B. Subspecialist response arrival times after Airway Emergency Pager System activation over the study duration. ns, no significant difference (p=0.053 by one-way ANOVA). C. The percentage of delayed subspecialist responses (arrival >5min after Airway Emergency Pager System activation) over the study duration. p=0.036 by linear regression.

A critical aspect of our program was rapid response and presence of subspecialists and equipment. Average time to first subspecialist arrival decreased, but not to statistical significance (Fig. 2B). We also tracked “delayed” subspecialist responses (>5 min), reasoning that it should take under 5 min to arrive at an acute event from anywhere in our hospital. Events with delayed subspecialist responses dropped dramatically (29±18% 2008-2012 vs 20±18%2013-2015 vs 9±11% 2016-2019, Fig. 2C, p=0.04 by linear regression). Delayed responses also decreased when analyzed on an annual basis (p=0.04, Supplemental Figure 1). Safety and assuredness afforded by consistently rapid responses were clinically meaningful.

Since program establishment, subspecialists and emergency equipment were present at every event. Specialized equipment usage has remained consistent over time (Supplemental Fig. 2A). Intubations were most frequently performed by ENT, although NICU or Anesthesia personnel also intubated successfully (Supplemental Fig. 2B). Few events (n=9, 5.7%) required management in an operating room. Review of these events highlighted a need for a 2.2 bronchoscope to be included with our airway emergency cart. There were 3 patient deaths (1.9% overall), 2 of which were clinically ascribed to airway issues. Multidisciplinary review of these events did not identify related system failures to necessitate changes in team organization or infrastructure.

The interdisciplinary collaboration promoted through our program was also critical in quickly and effectively adjusting procedures in response to the COVID-19 pandemic. Universal NICU admission screening, and standardized upper and lower respiratory tract sampling for intubated patients, enabled rapid identification of all COVID-19-positive patients. Given the likelihood of an ‘aerosol-generating procedure’ (i.e., intubation) at each emergency event, all staff wore fit-tested N95 masks, eye protection, and gowns and gloves donned and doffed per institutional personal protective equipment (PPE) policies. PPE and viral filters are now included in separate carts that were brought to all airway emergency events. Limited personnel were involved with COVID-19-positive airway interventions to limit exposure to these aerosol-generating procedures. Video laryngoscopic intubation was strongly preferred for these patients, as this allowed the greatest distance between the patient and providers.

## Discussion

Challenges associated with neonatal and infant airways are well described [2,3]. Our Airway Safety Program facilitated consistently prompt event management. As our program became standardized, we might have expected increased event frequency. Instead, the events remained consistent over time. We attributed this finding to prevalent use of video laryngoscopy in the past 2-3 years, which likely avoided some airway emergencies. Improved multidisciplinary communication may have also avoided some events. For example, the Airway Safety team was notified prior to certain events, such as the birth of infants with known airway malformations by scheduled induction or cesarean section in our Special Delivery Unit. Planned events were not included in this manuscript.

Our tracking system also enabled us to identify equipment most useful for successfully resolving airway emergencies. A 2.2 bronchoscope was not initially included with our specialized airway equipment. However, after program inception, this was clearly the dominant bronchoscope used in both diagnosis and airway access for most patients. We therefore include a 2.2 bronchoscope in our budget process and make it available during all emergency events.

This was an observational study based on a decade of experience. Airway emergency data were not systematically captured or evaluated prior to creation of this program. The relatively infrequent nature of these events limited our ability to demonstrate statistically significant differences between epochs, although the reliability of equipment and subspecialist personnel were clinically meaningful. The special patient population served in our quaternary NICU may limit the generalizability of our results, but we hope that our experience helps foster development of similar programs in other children’s hospitals.

Response times, equipment availability, and patient outcomes are key quantitative metrics with which to judge program success.

Our institution opened a Center for Fetal Diagnosis and Treatment in 1995. We have since experienced tremendous growth, evaluating and caring for more than 25,000 patients from 70 countries. Similarly, our institutional Neonatal Airway Safety Program, enacted in 2004, now handles 100-200 clinical airway management referrals per year. Many of these complex patients require expert airway management, necessitating a multidisciplinary team with collective experience treating fetuses and infants with congenital airway anomalies. Subspecialist cooperation and equipment availability that was streamlined through our Airway Safety Program experience has proven useful in managing these patients at delivery and throughout hospitalization.

Our program has created a safer care environment based on utilization of available expertise. Airway management programs are necessary at specialized birth centers and NICUs care for a growing number of infants with prenatally recognized airway anomalies. Multidisciplinary oversight and event reviews helped manage complications and systemic problems as they arose during program development, and facilitated rapid and effective adaptations in response to changing circumstances (e.g., the COVID-19 pandemic). Our hospital has now enacted a multidisciplinary Airway Emergency Response Committee to oversee and standardize equipment and responses in these challenging situations hospital-wide. We hope our experiences with our program will help others establish similar programs in other settings.

The COVID-19 pandemic allowed the multidisciplinary team to rethink PPE needs and our response practices to optimize safety and minimize the spread of aerosol generating contamination. Although robust data are not available for COVID-19-positive events, key changes in equipment and response practices described herein will be most important for other hospitals to implement a similar program.

## Conclusions

Establishment of an Airway Safety Program enhanced tracking and resource utilization in our quaternary children’s hospital. Subspecialist response times and equipment availability were streamlined as a result of this program. In many cases, expert users and specialized equipment were necessary to resolve life-threatening situations. Revising our protocols related to aerosol generating procedures in response to the COVID-19 pandemic was valuable for our institution. Similar programs could optimize management of clinically challenging airway emergency events in other hospital settings.

## Data Availability

De-identified summary data will be made available upon request.

## Acknowledgments

The authors are grateful to our respiratory therapists and nursing staff for diligently recording airway emergency events.

## Funding

This work was supported by the Children’s Hospital of Philadelphia Division of Neonatology and NIH 5T32HD043021 (CST).

## Conflicts of interest

The authors have no relevant conflicts of interest to disclose.

## Supplemental Information

### Supplemental Figures

**Supplemental Figure 1.**
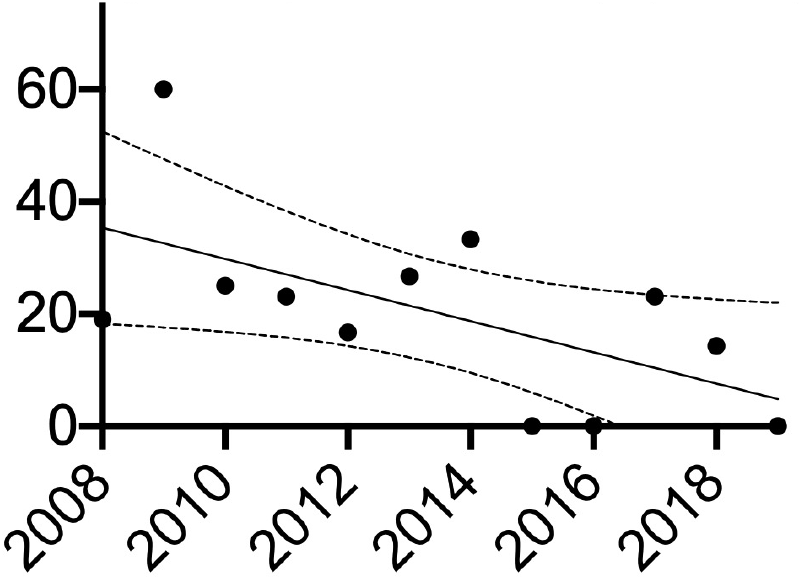
The percentage of delayed subspecialist responses (arrival >5min after Airway Emergency Pager System activation) over the study duration (linear regression slope -2.8, p=0.041).

**Supplemental Figure 2.**
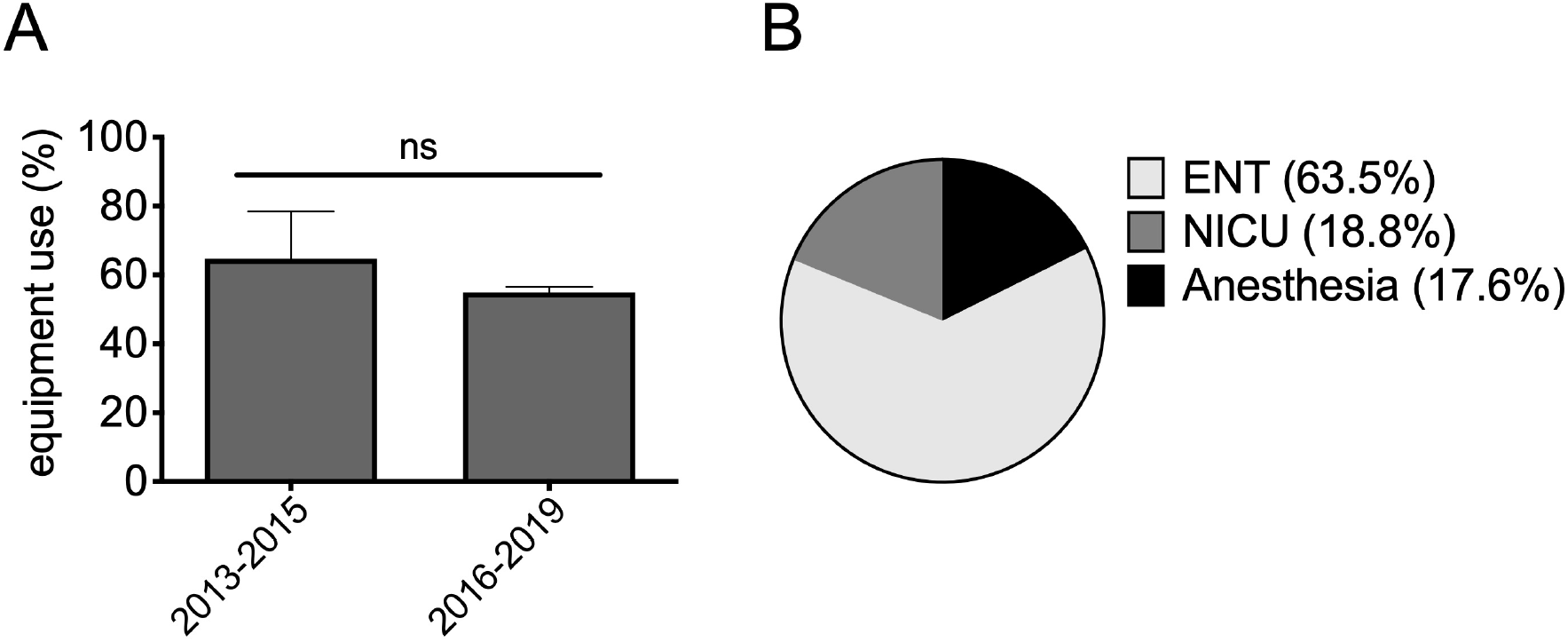
Percentage of events in which specialized equipment and personnel assisted in resolution of Airway Emergency Response events. A. The percentage (%) of events resolved with specialized equipment, such as Benjamin scopes, bronchoscopes, fiberoptic scopes, and/or other operating room equipment. ns, not statistically significant (p=0.44 by two-sided t-test). B. The percentage (%) of events at which the indicated specialist team intubated or otherwise resolved the Airway Emergency event.

## Notes

### Competing Interest Statement

The authors have declared no competing interest.

### Author Declarations

This study was deemed exempt from oversight by the Children's Hospital of Philadelphia Institutional Review Board (IRB 19-017149)

